# Community health workers for pandemic response: a rapid evidence synthesis

**DOI:** 10.1101/2020.04.28.20082586

**Authors:** S Bhaumik, S Moola, J Tyagi, D Nambiar, M Kakoti

## Abstract

**Introduction:** Coronavirus disease (COVID-19), a respiratory illness, first discovered in China in December 2019 has now spread to 213 countries or territories affecting millions across the globe. We received a request from National Health Systems Resource Centre, a public agency in India, for a Rapid Evidence Synthesis (RES) on community health workers (CHWs) for COVID-19 prevention and control.

**Methods:** We searched PubMed, websites of ministries, public agencies, multilateral institutions, COVID-19 resource aggregators and pre-prints (without language restrictions) for articles on the role, challenges and enablers for CHWs in pandemics. Two reviewers screened the records independently with a third reviewer resolving disagreements. One reviewer extracted data in a consensus data extraction form with another reviewer cross-checking it. A framework on CHW performance in primary healthcare not specific to pandemic was used to guide data extraction and narrative analysis.

**Results:** We retrieved 211 records and finally included 36 articles on the role, challenges and enablers for CHWs in pandemics. We found that CHWs play an important role in building awareness, countering stigma and maintaining essential primary healthcare service delivery. It is essential that CHWs are provided adequate Personal Protective Equipment (PPE) and appropriately trained in its usage in the early stages of the pandemic. Wide range of policies and guidance is required to ensure health systems functioning. A clear guidance for prioritizing essential activities, postponing non-essential ones and additional pandemic related activities is crucial. CHWs experience stigmatization, isolation and social exclusion. Psychosocial support, non-performance-based incentives, additional transport allowance, accommodation, child-support, awards and recognition programs have been used in previous pandemics.

We also created inventories of resources with guiding notes for quick utility by decision makers on guidelines for health workers (n=24), self-isolation in the community (n=10) and information, education and counselling materials on COVID-19 (n=16).

**Conclusions:** CHWs play a critical role in pandemics like COVID-19. It is important to ensure role clarity, training, supportive supervision, as well as their work satisfaction, health and well-being. There is a need for more implementation research on CHWs in pandemics like COVID-19.

**Summary Box:** *What is already known?:*

- COVID-19 is a pandemic which has now strained health systems across the world. Community health workers (CHWs) are being deployed in many countries for COVID-19 prevention and control. However, there is no evidence synthesis on the issue.

*What are the new findings?:*

- CHWs can play an important role in creating awareness, countering stigma, and maintaining essential primary healthcare delivery.
- Adequate provisions for personal protective equipment are essential for CHWs
- Governments should provide a clear guidance to CHWs for prioritizing essential activities, postponing non-essential ones and for additional pandemic related activities, while also ensuring their health and safety
- CHWs experienced stigmatization, isolation and were socially ostracized in previous pandemics. Psychosocial support, non-performance-based incentives, additional transport allowance, accommodation, child-support, awards and recognition programs have been used as enablers.

*What do the new findings imply?:*

- Roles of CHWs in a pandemic context must be clear. Apart from clear guidance adequate training and support should be provided. requiring adequate training and guidance is useful. Contact listing and identification should be done by separate trained cadre.
- Countries without pre-existing CHWs program must consider applicability of available evidence before investing in ambitious new CHW programs.
- There is a need for more implementation research on CHWs in pandemics like COVID-19.

## Introduction

The COVID-19 outbreak that originated in Wuhan City, China in December 2019 has emerged as a pandemic triggering a public health emergency worldwide. The disease now affects 213 countries or territories affecting millions across the globe. Several measures are being implemented by governments across the world to prevent and control COVID-19. Better health workforce utilisation and well-being is one of the key pillars governments across the world are dealing with to address the issue of health systems capacity. (1, 2) Community health workers (CHWs) constitute a significant frontline health workforce in many countries(3) and can potentially play an important role in control and prevention of COVID-19.

We received a request to examine the evidence to inform the potential role, enablers and barriers for CHWs (like Accredited Social health Activists or ASHAs in India) during COVID-19 prevention and control from the National Health Systems Resource Centre (NHSRC), India. The NHSRC is a public agency with the mandate to provide technical assistance to the National Health Mission. An inventory of resources which government decision makers and technical agencies could rapidly scan to help develop guidelines, standards of procedures, advisory notes and communication materials was also deemed to be a useful addendum by the requesters. We thus aimed to:

- understand key roles, issues, barriers and enablers for CHWs for pandemic response
- develop an inventory of resources that could be used to develop guidance, training manuals and IEC (information, education and communication) materials related to COVID-19 for CHWs

## Methods

### Approach for the study

Considering the urgency of evidence to inform decision making, we were provided a three-calendar day deadline by the requester. Based on an initial scoping, discussion with the requesters and the emergent nature of COVID-19, we expanded the scope to understand what can be learnt from previous pandemics, i.e. Severe Acute Respiratory Syndrome (SARS), Swine Flu, Ebola Virus Disease (EVD) and Middle East Respiratory Syndrome Coronavirus (MERS-CoV). We followed a Rapid Evidence Synthesis (RES) approach, wherein processes and methods of the traditional systematic review approach were tailored to ensure timeliness. (4) We took a broad scoping review approach for the RES to support the wide nature of decisions under consideration. There are no reporting guidelines specifically for rapid evidence synthesis, so we used the PRISMA checklist for scoping reviews.(5) The PRISMA checklist is presented in <u>Appendix 1</u>.

### Eligibility Criteria

We included studies which met the following criteria:

- Concept & context – studies or guidance related to roles, issues, challenges and enablers of CHWs during any phase of recent pandemics. The pandemics considered were SARS, Swine Flu, EVD, MERS-CoV & COVID 19.
- Types of participants- We did not use any specific definition of CHW recognising the fact that there are identified differently in different health systems across the world.
- Types of evidence sources – We included following types of evidence sources - primary research studies (of any design- qualitative or quantitative), any evidence synthesis, guidelines, training materials, advisories, standards of procedures, technical reports.

There were no language or date restrictions.

### Search methods for identification of records

We searched PubMed given the need to complete the RES in three days (Search Strategy in <u>Appendix</u> 2). We screened the reference list of all included studies to identify additional records and hand searched (MK, JT) 18 websites of different government, multinational agencies and COVID-19 resource aggregators and pre-prints <u>(Appendix</u> 3).

### Screening, Data collection and analysis

Two reviewers independently screened the titles and abstracts of studies for inclusion with a third author resolving consensus (SB, SM, JT). At the full text level, decision for inclusion or exclusion was done with the consensus of three authors (SB, SM, JT). Data from included studies was extracted using a pre-defined template by one reviewer (SM, JT, SB, DN) and cross-verified by at least one another reviewer. Cross-verification was not possible for the two French studies that were identified because only one of the reviewers knew the language (DN). No assessment of methodological quality of the studies was conducted as a part of methodological tailoring to expedite review process. (4)

We adapted an existing conceptual framework for assessment of CHW performance in primary health care (6) to guide data extraction and narrative synthesis. The framework looks at inputs, programmatic processes, CHW-level and community level outputs to improve health outcomes in the context of economic evaluation and equity, gender and accountability. The framework is not specific to pandemic response; for ease of reading and clarity, some components of the framework were merged, and issues related to multiple aspects of the framework mentioned without repetition.

## Results

### Search Results and Study Selection

We searched PubMed on 21 March 2020 and retrieved 211 records. Following title and abstract screening, full text articles were retrieved for 36 potentially relevant studies. Seven records were excluded following full text examination, leaving 29 studies for inclusion. An additional seven studies were identified by screening reference lists of the initial 29 studies that were included. Overall, 36 studies were included in the rapid review. A PRISMA flowchart for the same is presented in <u>Figure 1</u>. A list of studies excluded at full-text level is presented in <u>Appendix</u> 4.

**Figure 1.**
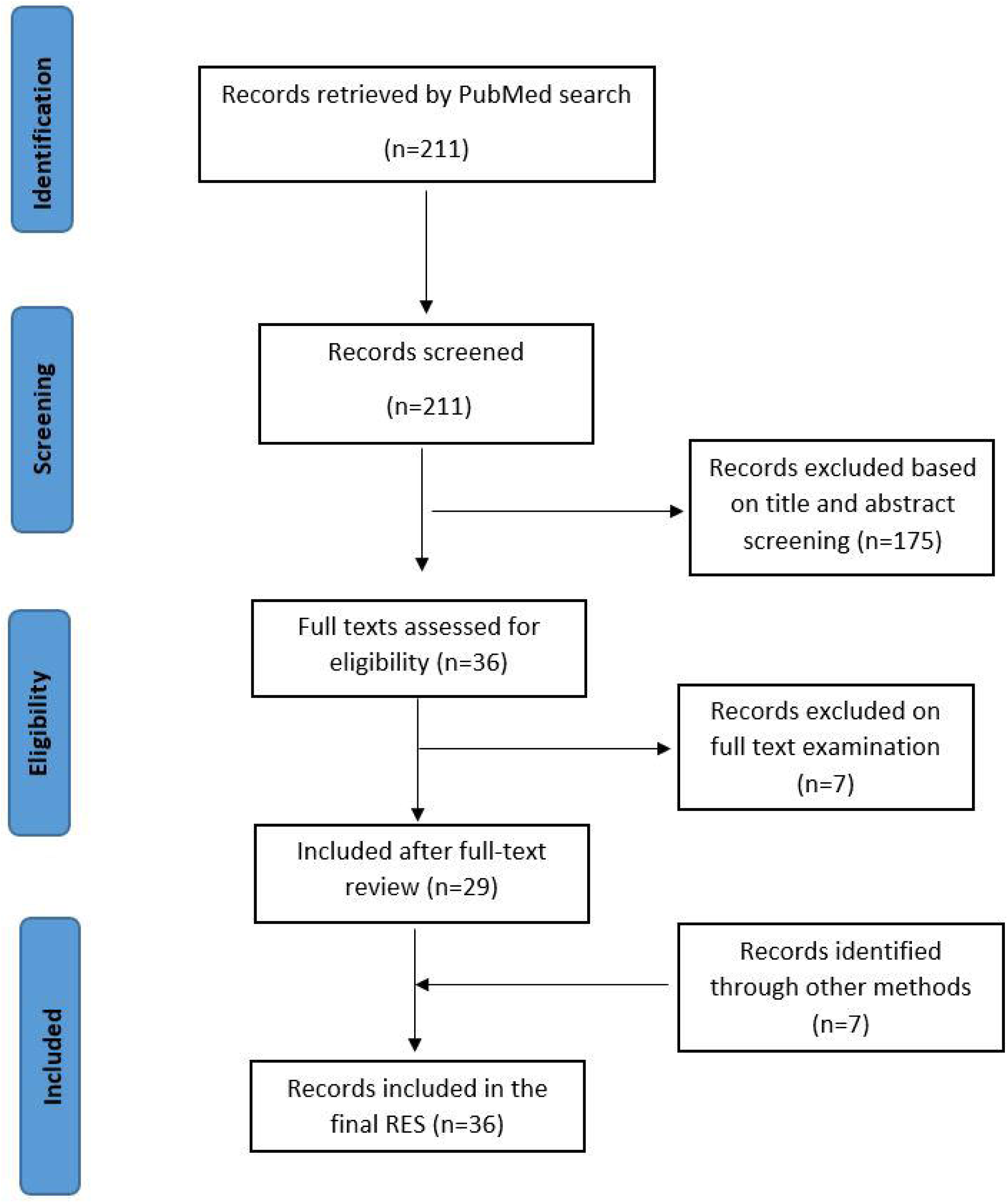

### Characteristics of included studies

A majority of the studies included in the review were qualitative (n=11) (7–17), followed by descriptive cross-sectional studies (n=9)(18–26), pre- and post-test studies (n=6) (27–32), literature reviews (n=3)(33–35), mixed-methods studies (n=2)(36, 37), guidelines (n=2)(38, 39), one systematic review(40), one randomised controlled trial (41), and one case report(42). Key characteristics of the included studies are presented in <u>Table 1</u>. Studies were conducted in Democratic Republic of Congo (DRC), Ghana, Guinea, Nigeria, Sierra Leone, Southern Sudan, Uganda, Bangladesh, Cambodia, Iceland, Laos, Thailand, UK, and Vietnam. The two guidelines were published by the World Health Organization (WHO).

**Table 1.**
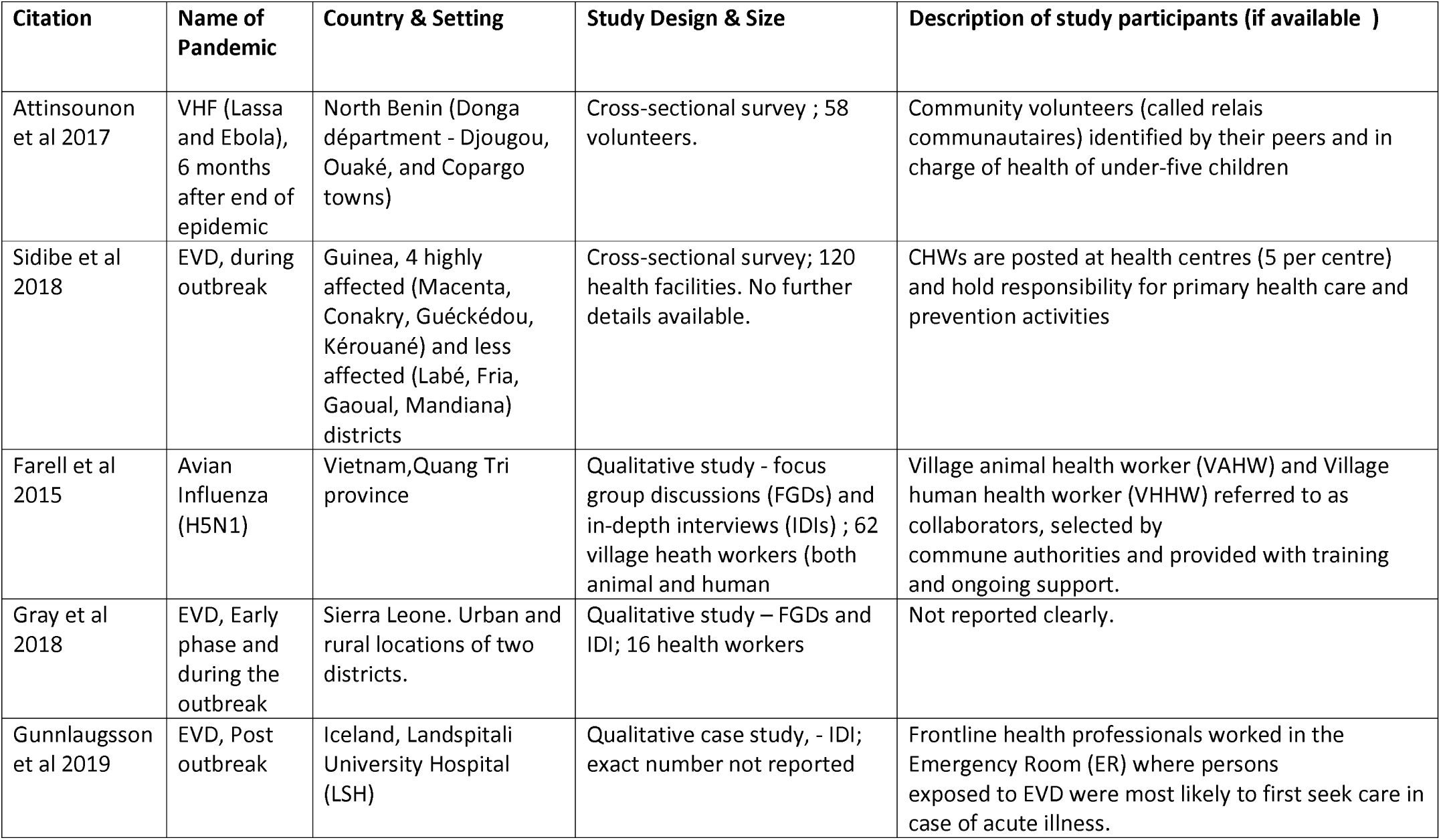
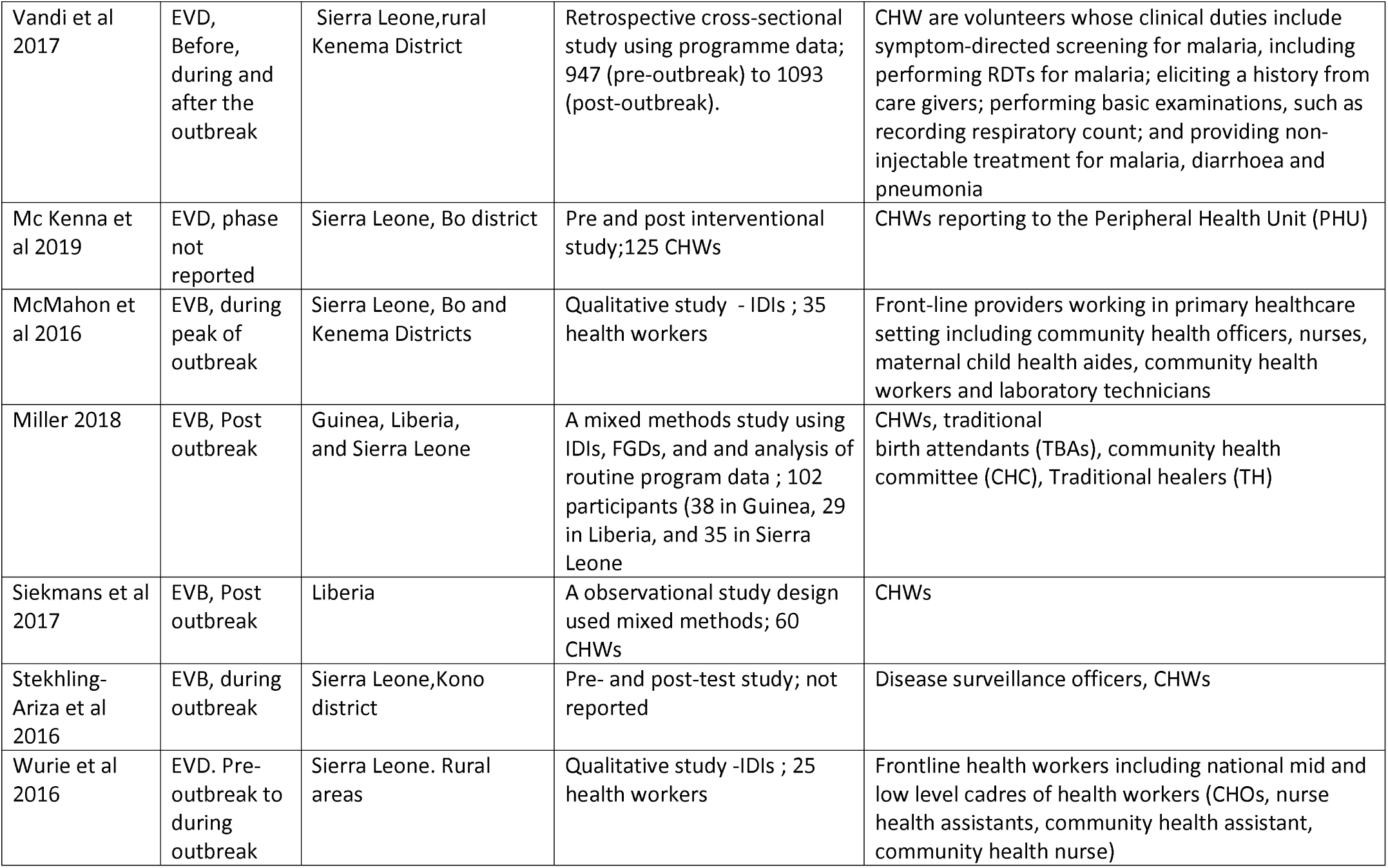
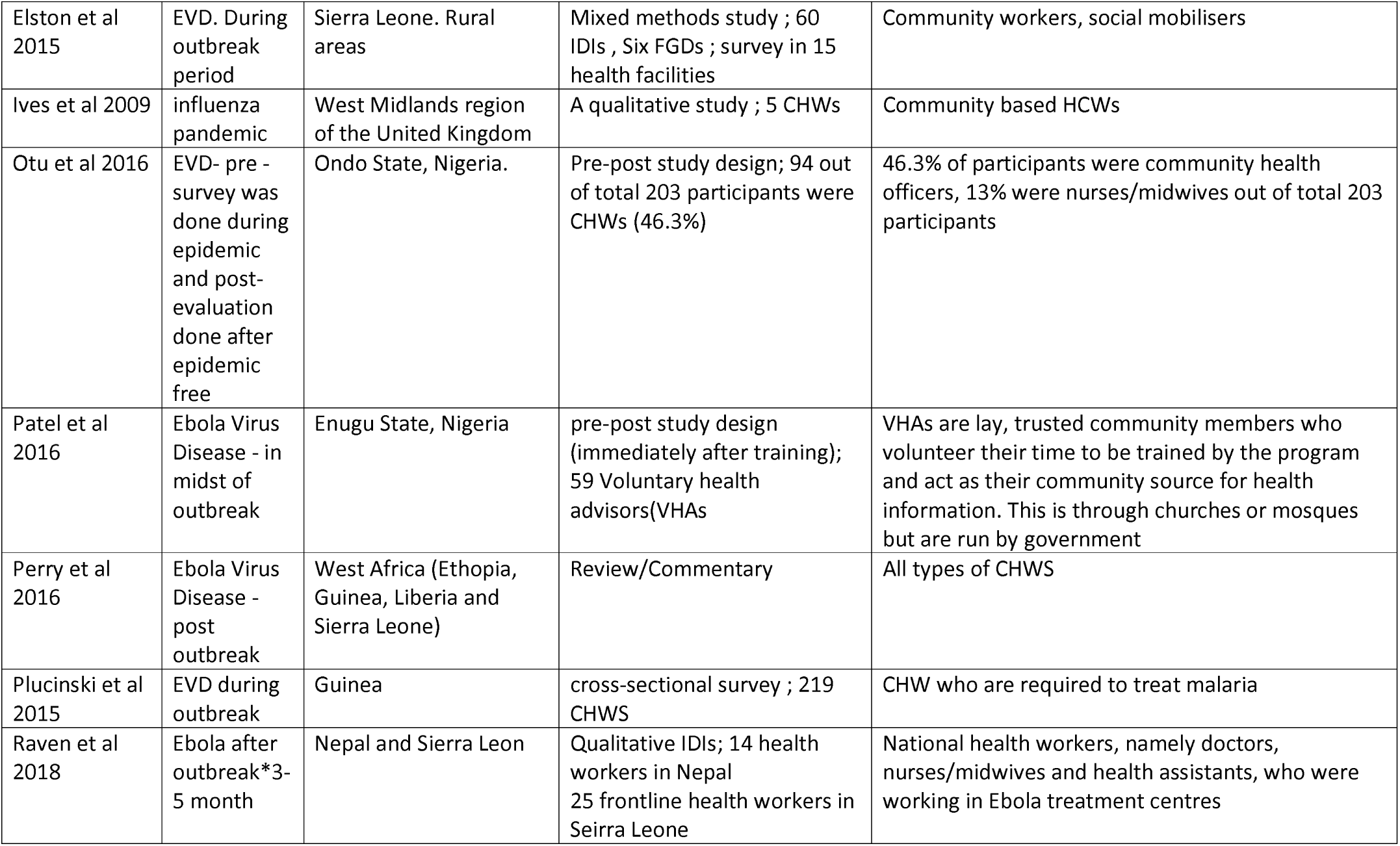
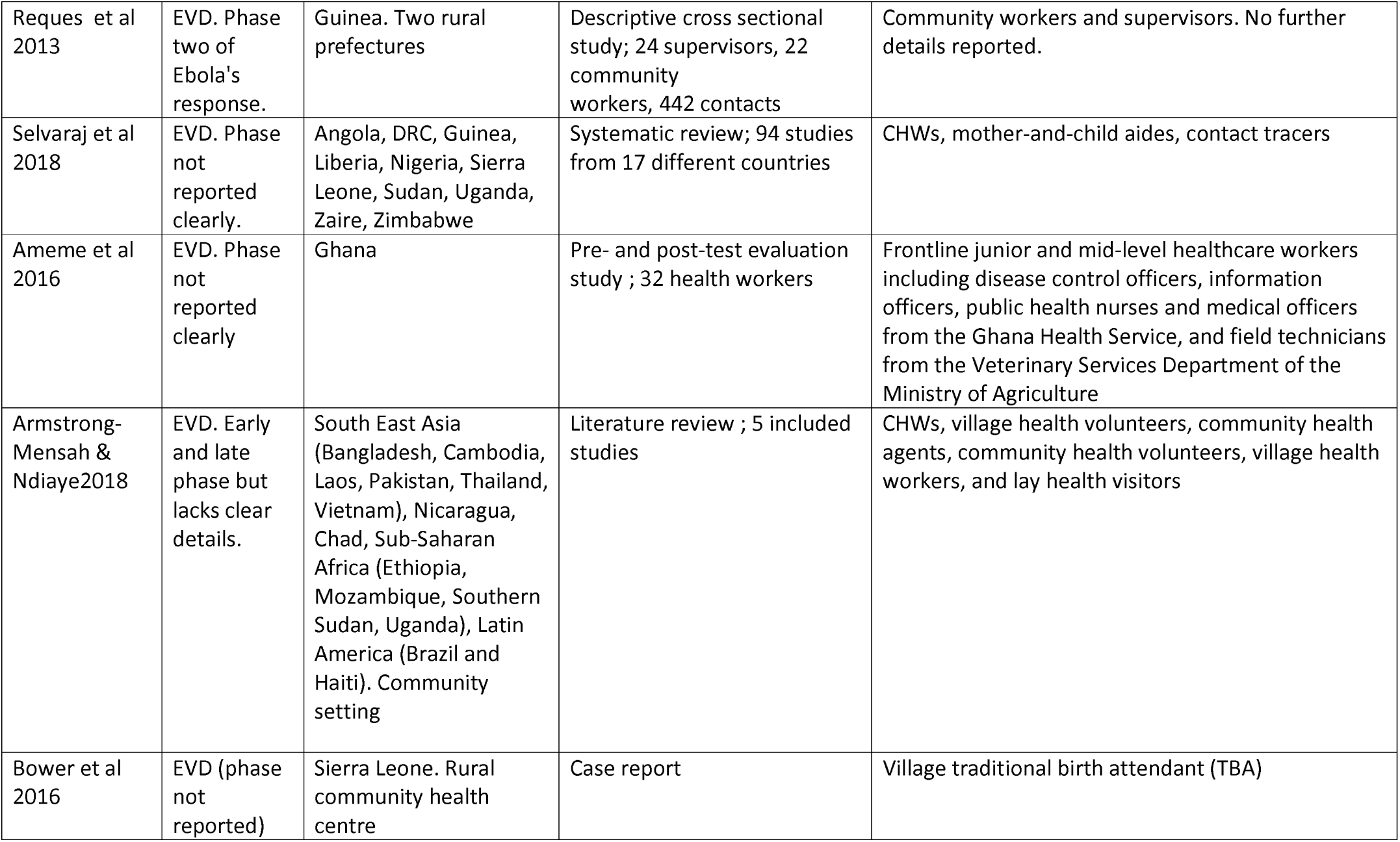
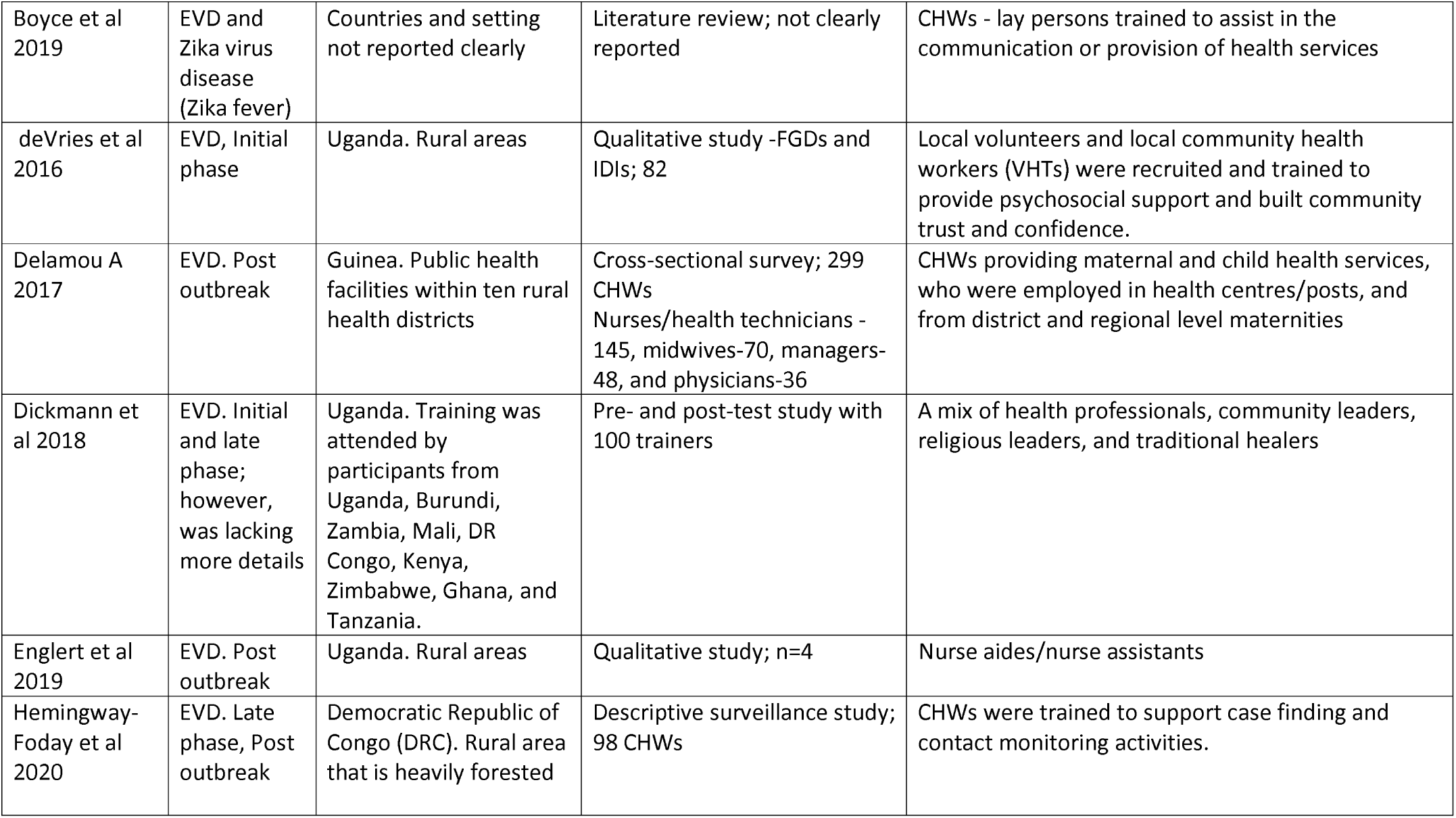
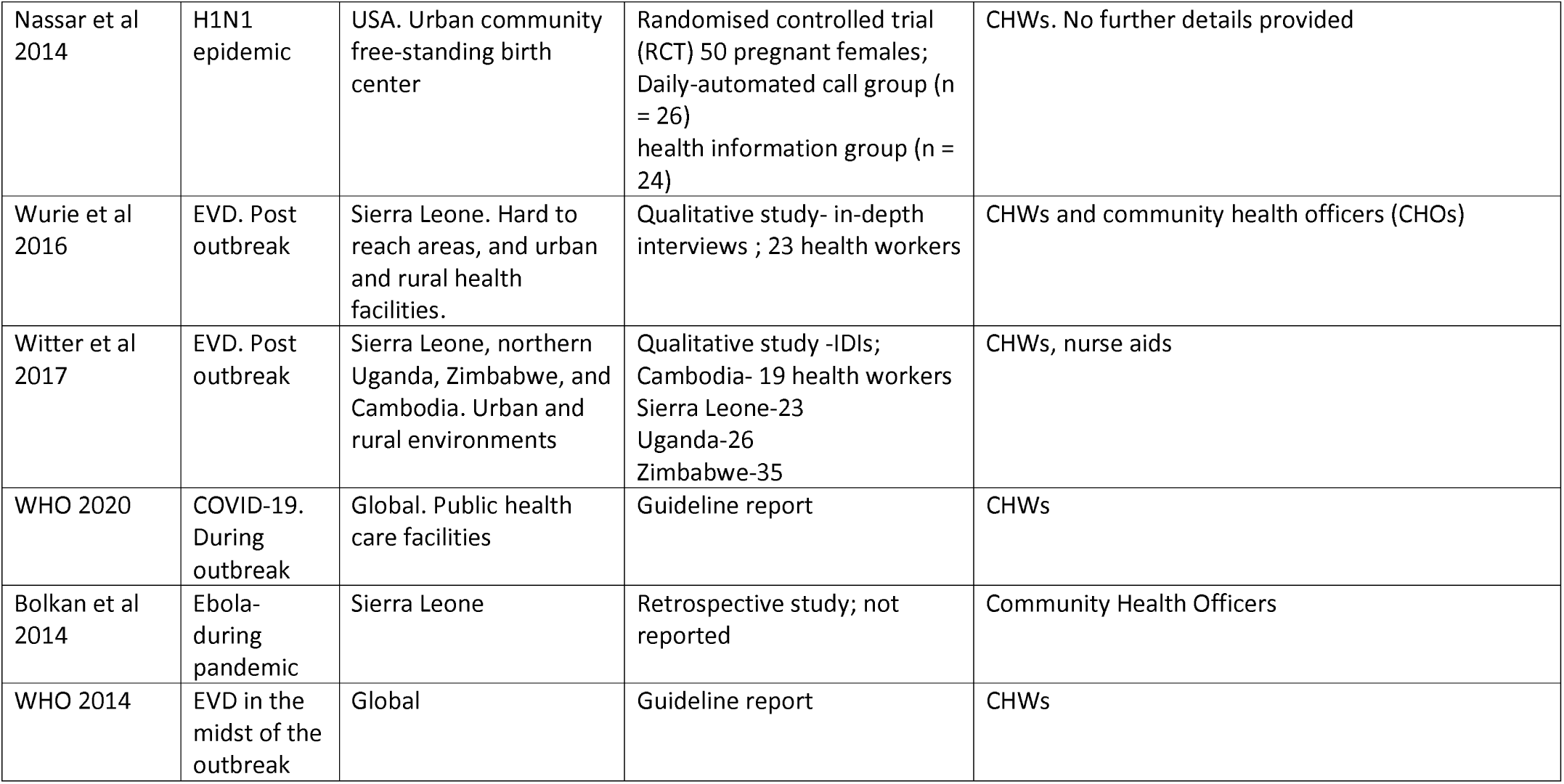

| Citation | Name of Pandemic | Country & Setting | Study Design & Size | Description of study participants (if available ) |
| --- | --- | --- | --- | --- |
| Attinsounon et al 2017 | VHF (Lassa and Ebola), 6 months after end of epidemic | North Benin (Donga département - Djougou, Ouaké, and Copargo towns) | Cross-sectional survey ; 58 volunteers. | Community volunteers (called relais communautaires) identified by their peers and in charge of health of under-five children |
| Sidibe et al 2018 | EVD, during outbreak | Guinea, 4 highly affected (Macenta, Conakry, Guéckédou, Kérouané) and less affected (Labé, Fria, Gaoual, Mandiana) districts | Cross-sectional survey; 120 health facilities. No further details available. | CHWs are posted at health centres (5 per centre) and hold responsibility for primary health care and prevention activities |
| Farell et al 2015 | Avian Influenza (H5N1) | Vietnam, Quang Tri province | Qualitative study - focus group discussions (FGDs) and in-depth interviews (IDIs) ; 62 village health workers (both animal and human | Village animal health worker (VAHW) and Village human health worker (VHHW) referred to as collaborators, selected by commune authorities and provided with training and ongoing support. |
| Gray et al 2018 | EVD, Early phase and during the outbreak | Sierra Leone. Urban and rural locations of two districts. | Qualitative study – FGDs and IDI; 16 health workers | Not reported clearly. |
| Gunnlaugsson et al 2019 | EVD, Post outbreak | Iceland, Landspítali University Hospital (LSH) | Qualitative case study, - IDI; exact number not reported | Frontline health professionals worked in the Emergency Room (ER) where persons exposed to EVD were most likely to first seek care in case of acute illness. |
| Vandi et al 2017 | EVD, Before, during and after the outbreak | Sierra Leone, rural Kenema District | Retrospective cross-sectional study using programme data; 947 (pre-outbreak) to 1093 (post-outbreak). | CHW are volunteers whose clinical duties include symptom-directed screening for malaria, including performing RDTs for malaria; eliciting a history from care givers; performing basic examinations, such as recording respiratory count; and providing non-injectable treatment for malaria, diarrhoea and pneumonia |
| Mc Kenna et al 2019 | EVD, phase not reported | Sierra Leone, Bo district | Pre and post interventional study; 125 CHWs | CHWs reporting to the Peripheral Health Unit (PHU) |
| McMahon et al 2016 | EVB, during peak of outbreak | Sierra Leone, Bo and Kenema Districts | Qualitative study - IDIs ; 35 health workers | Front-line providers working in primary healthcare setting including community health officers, nurses, maternal child health aides, community health workers and laboratory technicians |
| Miller 2018 | EVB, Post outbreak | Guinea, Liberia, and Sierra Leone | A mixed methods study using IDIs, FGDs, and analysis of routine program data ; 102 participants (38 in Guinea, 29 in Liberia, and 35 in Sierra Leone) | CHWs, traditional birth attendants (TBAs), community health committee (CHC), Traditional healers (TH) |
| Siekmans et al 2017 | EVB, Post outbreak | Liberia | A observational study design used mixed methods; 60 CHWs | CHWs |
| Stekhling-Ariza et al 2016 | EVB, during outbreak | Sierra Leone, Kono district | Pre- and post-test study; not reported | Disease surveillance officers, CHWs |
| Wurie et al 2016 | EVD. Pre-outbreak to during outbreak | Sierra Leone. Rural areas | Qualitative study -IDIs ; 25 health workers | Frontline health workers including national mid and low level cadres of health workers (CHOs, nurse health assistants, community health assistant, community health nurse) |
| Elston et al 2015 | EVD. During outbreak period | Sierra Leone. Rural areas | Mixed methods study ; 60 IDIs , Six FGDs ; survey in 15 health facilities | Community workers, social mobilisers |
| Ives et al 2009 | influenza pandemic | West Midlands region of the United Kingdom | A qualitative study ; 5 CHWs | Community based HCWs |
| Otu et al 2016 | EVD- pre - survey was done during epidemic and post-evaluation done after epidemic free | Ondo State, Nigeria. | Pre-post study design; 94 out of total 203 participants were CHWs (46.3%) | 46.3% of participants were community health officers, 13% were nurses/midwives out of total 203 participants |
| Patel et al 2016 | Ebola Virus Disease - in midst of outbreak | Enugu State, Nigeria | pre-post study design (immediately after training); 59 Voluntary health advisors(VHAs | VHAs are lay, trusted community members who volunteer their time to be trained by the program and act as their community source for health information. This is through churches or mosques but are run by government |
| Perry et al 2016 | Ebola Virus Disease - post outbreak | West Africa (Ethopia, Guinea, Liberia and Sierra Leone) | Review/Commentary | All types of CHWS |
| Plucinski et al 2015 | EVD during outbreak | Guinea | cross-sectional survey ; 219 CHWS | CHW who are required to treat malaria |
| Raven et al 2018 | Ebola after outbreak*3-5 month | Nepal and Sierra Leon | Qualitative IDIs; 14 health workers in Nepal<br>25 frontline health workers in Seirra Leone | National health workers, namely doctors, nurses/midwives and health assistants, who were working in Ebola treatment centres |
| Reques et al 2013 | EVD. Phase two of Ebola's response. | Guinea. Two rural prefectures | Descriptive cross sectional study; 24 supervisors, 22 community workers, 442 contacts | Community workers and supervisors. No further details reported. |
| Selvaraj et al 2018 | EVD. Phase not reported clearly. | Angola, DRC, Guinea, Liberia, Nigeria, Sierra Leone, Sudan, Uganda, Zaire, Zimbabwe | Systematic review; 94 studies from 17 different countries | CHWs, mother-and-child aides, contact tracers |
| Ameme et al 2016 | EVD. Phase not reported clearly | Ghana | Pre- and post-test evaluation study ; 32 health workers | Frontline junior and mid-level healthcare workers including disease control officers, information officers, public health nurses and medical officers from the Ghana Health Service, and field technicians from the Veterinary Services Department of the Ministry of Agriculture |
| Armstrong-Mensah & Ndiaye2018 | EVD. Early and late phase but lacks clear details. | South East Asia (Bangladesh, Cambodia, Laos, Pakistan, Thailand, Vietnam), Nicaragua, Chad, Sub-Saharan Africa (Ethiopia, Mozambique, Southern Sudan, Uganda), Latin America (Brazil and Haiti). Community setting | Literature review ; 5 included studies | CHWs, village health volunteers, community health agents, community health volunteers, village health workers, and lay health visitors |
| Bower et al 2016 | EVD (phase not reported) | Sierra Leone. Rural community health centre | Case report | Village traditional birth attendant (TBA) |
| Boyce et al 2019 | EVD and Zika virus disease (Zika fever) | Countries and setting not reported clearly | Literature review; not clearly reported | CHWs - lay persons trained to assist in the communication or provision of health services |
| deVries et al 2016 | EVD, Initial phase | Uganda. Rural areas | Qualitative study -FGDs and IDIs; 82 | Local volunteers and local community health workers (VHTs) were recruited and trained to provide psychosocial support and built community trust and confidence. |
| Delamou A 2017 | EVD. Post outbreak | Guinea. Public health facilities within ten rural health districts | Cross-sectional survey; 299 CHWs<br>Nurses/health technicians - 145, midwives-70, managers-48, and physicians-36 | CHWs providing maternal and child health services, who were employed in health centres/posts, and from district and regional level maternities |
| Dickmann et al 2018 | EVD. Initial and late phase; however, was lacking more details | Uganda. Training was attended by participants from Uganda, Burundi, Zambia, Mali, DR Congo, Kenya, Zimbabwe, Ghana, and Tanzania. | Pre- and post-test study with 100 trainers | A mix of health professionals, community leaders, religious leaders, and traditional healers |
| Englert et al 2019 | EVD. Post outbreak | Uganda. Rural areas | Qualitative study; n=4 | Nurse aides/nurse assistants |
| Hemingway-Foday et al 2020 | EVD. Late phase, Post outbreak | Democratic Republic of Congo (DRC). Rural area that is heavily forested | Descriptive surveillance study; 98 CHWs | CHWs were trained to support case finding and contact monitoring activities. |
| Nassar et al 2014 | H1N1 epidemic | USA. Urban community free-standing birth center | Randomised controlled trial (RCT) 50 pregnant females; Daily-automated call group (n = 26) health information group (n = 24) | CHWs. No further details provided |
| Wurie et al 2016 | EVD. Post outbreak | Sierra Leone. Hard to reach areas, and urban and rural health facilities. | Qualitative study- in-depth interviews ; 23 health workers | CHWs and community health officers (CHOs) |
| Witter et al 2017 | EVD. Post outbreak | Sierra Leone, northern Uganda, Zimbabwe, and Cambodia. Urban and rural environments | Qualitative study -IDIs; Cambodia- 19 health workers<br>Sierra Leone-23<br>Uganda-26<br>Zimbabwe-35 | CHWs, nurse aids |
| WHO 2020 | COVID-19. During outbreak | Global. Public health care facilities | Guideline report | CHWs |
| Bolkan et al 2014 | Ebola- during pandemic | Sierra Leone | Retrospective study; not reported | Community Health Officers |
| WHO 2014 | EVD in the midst of the outbreak | Global | Guideline report | CHWs |

### Summary of findings on role, issues, enablers and barriers for CHWs in pandemic response

The included studies examined the role of CHWs in disease outbreak situations, in addition to exploring the barriers and enablers of working in pandemic situations. Overall, the studies reported that CHWs had a role to play in terms of community engagement (mostly in rural areas) and creating awareness during pandemic responses. The key findings from the included studies are presented as per the elements of the conceptual framework.

### Input in CHW programs during pandemic responses

Myriad inputs and planning were required to ensure effective pandemic response through CHWs. These are summarised in sub-domains below:

- **Policies related to CHW task/roles** - Policies and guidance related to specific roles and tasks for CHWs were reported in many studies.(9, 21, 30, 34, 37, 38) These policies largely aimed to distinguish tasks as essential (routine activities that need to be continued but with modifications for decreased transmission risk), non-essential (nonessential activities that could be postponed) and additional (activities that need to be carried out additionally owing to the pandemic) to bring accountability and enhance trust in health systems. CHWs have been involved in various pandemic control activities, including community engagement, community sensitisation, awareness and promotion and use of appropriate preventive practices. These were reported to be crucial for the purpose of bringing accountability and building trust in health systems. The World Health Organization’s guidance on contact tracing recommends contact identification and listing to be conducted by a trained epidemiologist or surveillance officer, while contact follow-up might be done through CHWs.(38) However, polices to involve CHWs in some aspects of contract tracing had been developed and implemented during EVD. (21, 22)
- **Logistics & funding for CHW programs**. Disruptions in drug and equipment supplies were common during pandemics and as such, policies and mechanisms to address them from an early phase of the outbreak were reported to be important. (5, 10, 16) As such this might be expected along with shortfalls in routine supportive supervision methods. Sustained investments in CHWs is required during pandemic response including revisiting financial investments (discussed subsequently) for optimal outcomes.(34)
- **Governance and Stakeholders** – A wide range of stakeholders in the community need to be engaged. The evidence base is mostly from countries with existing CHW programs where they were additionally engaged for COVID-19 activities and has been discussed in a subsequent domain
- **Information Management Systems for CHWs in Pandemics** - Operational support using information management systems and digital health technology had been used for supporting and monitoring CHW programs. Honesty, transparency of communication and reciprocity of information with CHWs was noted to be beneficial (9, 14, 21).

### Programmatic Processes & Community health systems for CHWs in pandemic response

Programmatic process and health systems were crucial for effective service delivery by CHWs during pandemics.

#### Supportive systems, development, competency and well-being for CHWs

CHWs and their supervisors required appropriate training and supportive supervision for community sensitisation, awareness and risk communication during pandemic outbreaks and the lack of these was noted as a significant barrier to effective service delivery. (7, 9, 13, 16–18, 20, 21, 25, 29, 32, 34, 37, 38, 40) In most of these studies, it was reported that appropriate training in skills and knowledge related to disease outbreaks led to an improvement in awareness of the disease, screening and reporting. In qualitative studies, it was reported that there was a change in perceptions of CHWs, leading to more proactive involvement in disease prevention and control. Appropriate training helped CHWs overcome fear and become more confident about providing and delivering care and helped them deal with a broad range of challenges. Transparency in communication that included sharing of information from supervisors motivated CHWs to be involved, and mitigated the fear frequently encountered during pandemics.

CHWs were reported to be at an increased risk of exposure due to lack of or insufficient or incorrect usage of personal protective equipment (PPE).(10, 40) In previous disease outbreaks, training sessions on pandemic preparedness were focussed on protocols, and the use of appropriate and adequate PPE, during patient care. There is some evidence that training and availability of PPE resulted in CHWs being more confident to cope with managing the disease outbreak.(10, 13, 16, 17, 20, 40, 42)

During previous pandemics, CHWs have faced stigmatisation, isolation and in some cases were even socially ostracized.(15, 33, 37) CHWs used several coping strategies including trying to isolate themselves to protect their loved ones, finding renewed purpose in continuing to serve the community, and turning to religion to guide strength.(15) Peer and family support (praying together before work or through social-medial platform like WhatsApp) were reported to enhance CHW well-being.(15) Further, workshops that provided emotional support and ways to deal with the social stigma helped CHWs cope with seeing patients and colleagues dying from the disease. Studies reported that improved working conditions, provision of housing allowance, equal training opportunities, transportation allowance, and improved salaries (paid on time and for a broad range of services), awards in high-profile public events contributed to better recruitment and retention of CHWs during pandemics.(13, 15, 16, 33, 34)

#### Support from community-based groups for community access and community-centred care

Community engagement and sensitisation were reported to address knowledge gaps related to disease outbreak and discourage discrimination and stigmatisation towards CHWs. (15, 33) Coercive laws against community members initiating discrimination and stigmatisation towards CHWs were not reported. Information sharing in the community as well as engagement of community leaders were reported to help build trust in CHWs, leading to better access as well as satisfactory experience in care received.(8, 28, 33, 35, 37) Community elders and village heads should be engaged and involved to rebuild trust, leading to an increased use of services.

### Community level outputs and health outcomes of CHW-led pandemic response

A study noted that the social standing (in terms of social status) of CHWs empowered them in terms of their participation in pandemic responses.(34) Community health policies for social mobilisation and community engagement strategies were found to build trust and increase utilisation of services, which were found to be useful in pandemic response.(8, 15, 33, 37) Engaging CHWs in the early phases of outbreaks potentially improved overall response procedures and adaptive resilience.(34) It was reported that CHW-led programs led to improved knowledge and awareness among farmers leading to improved motivation for reporting of avian influenza.(7) In another study, it was noted that reporting for malaria by CHWs improved from 59% (pre-outbreak) to 95% (during outbreak) and 98% (post-outbreak) owing to “sustained investments in CHWs through provision of incentives, supervision and provision of adequate reporting tools”.(22) In general, training of CHWs led to improved knowledge, service quality and well-being, as has been noted in preceding sections. One study from rural Uganda reported that CHW-delivered psychosocial support to build community trust and confidence was often not available for disadvantaged and poor families owing to lack of accountability measures.(12) As such it is important to build adequate governance and reporting structures and use an equity focussed approach in programming.

### Equity, Gender, Accountability and Economic Evaluations

We did not find any economic evaluations related to CHWs in pandemic responses. Issues around equity, gender and accountability were seldom studied, including in relation to the populations to whom CHWs provided their services. Specific equity focussed studies were scarce but female CHWs had a different set of challenges, which has been discussed in an earlier section pertaining to CHW support and well-being.

### Summary of inventory of resources to guide resource development

We created three inventories to guide and expedite resource development. The first inventory (<u>Appendix 5</u>) compiled 24 documents. Apart from guidance documents useful for health workers (not just CHWs) during pandemics the inventory also included resources for ensuring occupational safety and well-being and listing actions for local health departments and sub-national planning.

The second inventory included 10 guidelines <u>(Appendix 6</u>) and advisories on self-isolation practices which have been released by various government health departments to control COVID-19. They provide instructions for the 14-day home-quarantine in non-healthcare settings i.e., at home for those with close contact to COVID-19 cases, inbound travellers from affected areas, and people sharing the same home with a suspect or positive COVID-19 person.

The third inventory consisted of list of 16 different IEC resources <u>(Appendix 7</u>). They address frequently asked questions, list COVID-19 myths and facts, Dos and Don’ts for the general public, and specific communication messages for high-risk population and individuals showing symptoms.

## Discussion

### Summary of main results

The studies included in the review reported that CHWs had insufficient knowledge about disease outbreaks; however, showed positive perceptions of the prevention and control of disease transmission. The evidence from our RES pertains to settings where well-established CHW programs exist before a pandemic. The findings of this review suggest significant knowledge gaps between the amount of information available on disease outbreaks and the depth of knowledge among CHWs, particularly about the mode of transmission, and the use preventive and control measures, such as the use of PPE measures.

### Completeness and applicability of the evidence

In terms of the strengths of this review, a robust and transparent search strategy was utilised to identify relevant studies of interest. Broadening the scope to include multiple recent pandemics enabled us to get more meaningful evidence from a broader but relevant literature to inform decision making. We searched only one database, but we had extensively used other methods to identify other research. The identification of seven studies by other methods demonstrate the robust application and utility of the approach. We acknowledge that a few more studies on the topic might have not been retrieved but contend this would not have changed the results of the study majorly.

### Implications for policy and practice

Evidence from our RES indicates that in countries with well-established CHW programs, this cadre can play a very important role in increasing awareness and countering stigma. CHWs should continue to maintain essential primary healthcare service delivery, with a separate trained cadre being used for contact identification and listing. This is crucial considering the larger body of evidence from previous pandemics showing consequences on population health owing to disruption in routine service delivery.(43–45) Disruption in supply-chain, logistics and supportive supervision is expected and a clear guidance (with requisite training) for prioritising essential activities, postponing non-essential ones is crucial. The CHWs should be provided adequate PPE and be appropriately trained in its usage in the early phases of the pandemic.

During previous pandemics, CHWs experienced stigmatisation, isolation and were socially ostracised, and the same might be expected during COVID-19. The government should thus make provisions for psychosocial support, move to a non-performance-based incentive model or regularisation of staff and institute awards and recognition programs. Provisions for additional transport allowance, accommodation, and child-support should also be considered owing to the need to protect families of CHWs from infection. As most CHWs are women, they are subject to gender norms that on the one hand may increase infection risk (domestic work) as well as other gendered vulnerabilities and risks (like domestic violence)(46). Sustained investment together with governance and information systems support is important. Further, as argued elsewhere, CHWs are part of a larger ecosystem of health system wherein ensuring trust and accountability is essentially and more so during a pandemic(44).

Implications for introducing new CHW programs in the midst of a pandemic, as is being planned by the United Kingdom(47) is not well understood, but considerable challenges might be expected considering the larger body of literature around community trust, and systems integration being critical for success.(48–51)

### Policy Impact and Stakeholder Engagement

We submitted the key policy considerations along with a shorter version of the RES to the requester and released it publicly on our institutional website considering the immediacy of decision making. The RES has been used for development of training, recommendation adoption in Odisha, a state in eastern India and been referred to in WHO policy note on violence against women (46) during COVID-19. The inventories have also been used extensively for preparation of checklists for rural and urban primary health centre preparedness to tackle COVID-19. (52, 53) Our detailed experience in relation to stakeholder engagement for uptake of RES will be captured in a separate paper.

### Implications for research

We took a broad scoping approach in the RES to understand issues around CHWs in pandemic response. Conduct of research related to health workers during pandemics is extremely challenging and as such, high-quality evidence from trials may be neither feasible nor ethical. However, the use of cross-sectional surveys, before-and-after studies, time-series studies, and qualitative study designs is feasible. Implementation research related to key pandemic response functions like contact tracing, community awareness and anti-stigma programs is crucial to understand issues related to human resources for health and better pandemic control. Equity, gender and accountability dimensions together with economic evaluations are major gap in the literature that needs to be studied.

## Conclusion

In conclusion and based on our findings, CHWs play an important role in the prevention and control of pandemics like COVID-19. This is relevant and feasible in countries which already have existing CHW programs. However, there is a need for role clarity, further training, and regulation to ensure preparedness of CHWs in pandemic-like situations. The use of CHWs in countries without pre-existing CHWs programs may be challenging considering the health systems re-orientation needs and the lack of established community relationships. Applicability of available evidence on CHWs for pandemic response should be considered by such countries before embarking on ambitious CHW programs amid a public health crisis.

## Data Availability

Data related to study is presented open access in appendices

## Acknowledgment

Dr. Rajani R. Ved and Dr. Neha Dumka from National Health Systems Resource Centre for inputs on scope of the RES to enhance utility in government decision making.

